# Correlation between renal function and peripapillary choroidal thickness in treatment naïve diabetic eyes using SS-OCT

**DOI:** 10.1101/2020.02.18.20024760

**Authors:** Sen Liu, Wei Wang, Yan Tan, Miao He, Lanhua Wang, Yuting Li, Wenyong Huang

## Abstract

**Purpose:** To investigate the association between the estimated glomerular filtration rate (eGFR) and peripapillary choroidal thickness (pCT) and retinal nerve fibre layer (pRNFL) thickness in diabetic patients by using swept-source optical coherence tomography (SS-OCT).

**Methods:** Ocular treatment-naïve patients with type 2 diabetes mellitus registered in the community health system in Guangzhou, China were recruited to participate in this prospective cross-sectional study. The eGFR was determined using the Xiangya formula, and the renal function was categorized into non-chronic kidney disease (non-CKD), mild CKD, and moderate to severe CKD (MS-CKD) according to the guidelines. The pCT and pRNFL thicknesses at 12 o’clock were obtained using a SS-OCT by a circular scan with a diameter of 3.4 mm centring on the optic nerve head, and the data from only one eye in each patient were used.

**Results:** This study included 1,408 diabetic patients, with a mean age of 64.4±7.8 years. The average pCT decreased with renal function deterioration, with 126.0 μm ± 58.0 μm for non-CKD, 112.0 μm ± 51.2 μm for mild CKD and 71.0μm ± 22.9 μm for MS-CKD, respectively (P<0.001). The pCT was found to be significantly thinner in CKD patients in all quadrantes (P < 0.05 in all regions) with the exception of the inferior quadrant, and the average pCT was positively correlated with eGFR (β = 0.3, 95%CI = 0.0 to 0.6, p = 0.021) after making adjustments for other factors. The pRNFL thickness in the nasal quadrant was significantly reduced in patients with CKD, and pRNFL thickness was positively correlated with eGFR (β = 0.1, 95%CI = 0.0-0.2, p = 0.009) after adjusting for other factors.

**Conclusion:** Impaired renal function was associated with a reduction of pCT and pRNFL thickness in patients with type 2 diabetes. The measurement of pCT and pRNFL may provide additional information for predicting renal impairment.

## Introduction

The disease and economic burden of diabetes mellitus (DM) has been increasing consistently around the world. In 2013, there were 382 million adults with DM, and the number is estimated to reach 592 million by 2035.^1^ Diabetic retinopathy (DR) and diabetic nephropathy (DN) are the most essential microvascular complications.^2^ The former is the leading cause of blindness in the working age population in industrialized countries, and the latter is the leading cause of end-stage renal diseases. Since DR and DN lack obvious clinical manifestations in their early stages, these patients were frequently underdiagnosed. DR has been screened in many countries using retinal photography, but the evaluation of renal function in diabetic patients remains challenging.^3^ Serum creatinine (Scr) and microalbuminuria are the common indicators for renal function, but the determination relies on invasive blood sample collection and complicated laboratory tests. In addition, the microalbuminuria as a screening maker can only be suggested when diabetic kidney damage has severely progressed.^4^ The accuracy is also limited due to sample preservation and contamination caused by variations in screening environments.

The retina can be easily accessed, and imaging can be conducted in vivo. DR and DN share common vascular risk factors and pathogenesis mechanisms. Measuring retinal parameters using non-invasive methods may help elucidate the early mechanisms of microvascular complications of DM, and it may also identify individuals at high risk of nephropathy in early stages of diabetes.^5^ Over past decades, a number of studies have confirmed the close association between DR signs, retinal vascular calibre and DN risk by using retinal photography.^6-8^

The retina nerve fibre layer (RNFL) and choroid are important structure of the retina, with the former providing information related to nerve damage and the latter receiving around 95% of the total blood flow to the eye, which provides nutrition for the outer layer of the retina. Recent studies have suggested that peripapillary RNFL loss is associated with impaired renal function in diabetic patients.^9^ There studies demonstrated the thinning of the choroid associated with DR, suggesting the choroid may be involved in DR pathogenesis.^10-13^ However, since a majority of previous studies of choroid focused on macula, little information was available on the alteration of choroid in DM.^10-13^ In addition, the influence of renal function on peripapillary choroidal thickness remains unclear. To fill the knowledge gaps in this field of study, this cross-sectional study of diabetic patients was performed to investigate the association of eGFR with pCT and pRNFL thicknesses in diabetic patients by using an innovative SS-OCT method.

## Materials and Methods

### Participants

This was a prospective cross-sectional study and performed at the Zhongshan Ophthalmic Centre (ZOC), Sun Yat-sen University, China. This study was approved by the Institute Ethics Committee of ZOC, and performed in accordance with the tenets of the Helsinki Declaration. Signe informed consent forms were obtained from all participants. Ocular treatment-naïve patients with type 2 diabetes aged 40 or greater were recruited from the community health system in Guangzhou, China. The participants were excluded if any of the following conditions were present: cylinder degree of −12 diopters (D) or more, an axial length of 30 mm or more, a best corrected visual acuity (BCVA) of 0.1 or less (Snellen chart), an intraocular pressure (IOP) of 21 mmHg or more, a C/D ratio of 0.5 or more, an inter-eye asymmetry 0.2 or more, a history of ocular disease (except for light cataract or refractive errors) or trauma, a history of ocular laser or surgical interventions or a history of other systemic diseases, such as myocardial infarction and stroke.

### General information and laboratory parameters

General information, including age, sex, length of time the patience has had diabetes, other systemic chronic diseases and risk factor, was collected via questionnaires. The weight, height, systolic blood pressure (SBP) and diastolic blood pressure (DBP) were measured by a nurse using standard procedures. Blood and urine samples were obtained from all patients, and the following parameters were analysed: serum creatinine (Scr), haemoglobin A1c (HbA1c), total cholesterol (TC), high-density lipoprotein cholesterol (HDL-C), low-density lipoprotein cholesterol (LDL-C), triglyceride (TG) and microalbuminuria. Recent studies have demonstrated that the traditional eGFR equation displayed limited accuracy in the Chinese population, and the latest Xiangya formula displayed the highest accuracy.^14^ In this study, the eGFR was calculated based on the Xiangya formula. The patients were categorized into 3 groups: the non-chronic kidney disease (non-CKD) group for patients with an eGFR of 90ml/min/1.73m^2^ or more, the mild-CKD group for patients with an eGFR of 60 ml/min/1.73m^2^ to 89 ml/min/1.73m^2^ and the moderate to severe-CKD (MS-CKD) group for patients with an eGFR less than 60 ml/min/1.73m^2^.

### Ocular examination

Comprehensive ocular examinations were conducted for all participants. The anterior and posterior segments were evaluated by slit-lamp biomicroscopy and ophthalmoscopy. The evaluations of best-corrected visual acuity (BCVA) and intraocular pressure (IOP) were regularly measured. Ocular biometric parameters were obtained using optical low-coherence reflectometry (Lenstar LS900; Haag-Streit AG, Koeniz, Switzerland). Auto refraction was measured by an autorefractor (KR8800; Topcon, Tokyo, Japan) after pupil dilation.

### SS-OCT Imaging

An SS-OCT (DRI OCT-2 Triton; Topcon, Tokyo, Japan) instrument was used to obtain high-definition retina and choroid images. The SS-OCT parameters included a wavelength of the SS-OCT light source = 1,050 nm, a scan rate = 100,000 A-scans per second, a depth of the scan window = 2.6 mm, an axial resolution = 8 μm and a transverse resolution = 10 μm. Segmentation of different layers on the OCT images and construction of topographic maps were automatically performed by built-in software. The segmentation was carefully inspected and confirmed by the same OCT technician who was blind to the refractive status of the participants, and manual correction was performed when the software misjudged the borderline of each layer. The peripapillary region was scanned using a 360 degree, 3.4-mm-diameter circle that was centred on the optic disc. Scans were centred using an internal fixation. The pCT was automatically measured as the perpendicular distance between the outer portion of the hyperreflective line corresponding to the retinal pigmented epithelium (RPE) and the hyporeflective line or margin corresponding to the choroidal-scleral interface. The pRNFL thickness was defined as the vertical distance between the internal limiting membrane (ILM) and the interface between the retinal nerve fibre layer and the retinal ganglion cell layer. The pCT and pRNFL was displayed in 12 sectors (12 o’ clock). The average pCT/pRNFL and pCT/pRNFL values in the superior, inferior, temporal and nasal quadrants were also calculated for statistical analysis.

### Statistical analyses

The Kolmogorov-Smirnov test was performed to verify normal distribution. When normality was confirmed, the analysis of variance (ANOVA) test was conducted to evaluate the inter-group difference of demographic, systemic and ocular parameters. If there were statistical differences, a pairwise comparison between groups was conducted by the Tukey method. The Fisher’s exact test was used for categorical variables. Bivariate scatter plots were generated to display the potential factors affecting pCT and pRNFL thicknesses. Variables with a P value <0.10 in univariate analyses were included in the multivariate analysis. Univariate and multivariate linear regression analysis were used to assess the associations between pCT/pRNFL thicknesses and renal function. When analysing the relationship between renal function and pRNFL/pCT thicknesses, renal function as an ordinal category variable was considered to be the independent variable, non-CKD was the dummy variable, and pRNFL and pCT thickness was the dependent variable for linear regression analysis. Potential confounding factors were adjusted, with model 1 adjusted for age and sex, and model 2 adjusted for age, sex, duration of diabetes, systolic blood pressure, BMI, triglyceride, cholesterol, HbA1c, insulin use, axial length and the presence of DR. First, when analysing the correlation between eGFR and pCT/pRNFL thickness, univariate linear regression analysis was used. Further, multiple linear regression adjusted for potential confounders was used. A P value less than 0.05 was considered significant. A P value of <0.05 was considered statistically significant. All analyses were performed using Stata, version 14.0 (Stata Corporation, College Station, TX, USA).

## Results

### Demographic and clinical features

A total of 1,408 patients who met the inclusion/exclusion criteria were included in the final statistical analysis. The basic demographic and clinical features are shown in Table 1. The mean age was 64.4 ± 7.8 years, with 774 (58.33%) female and 634 (41.67%) male patients. The mean age was 59.1 ± 8.9 years for the non-CKD group, 64.7 ± 7.5 years for the mild-CKD group and 71.3 ± 5.6 years for the MS-CKD group. The three groups differed significantly in age (p<0.001), course of diabetes (p<0.001), serum creatinine (p<0.001), triglyceride (p=0.015), serum uric acid (p<0.001), C-reactive protein (p=0.002), microalbuminuria (p<0.001) and best corrected visual acuity (p=0.004), while other variables included in the study showed no significant difference among the three groups.

**Table 1.** Demographic and clinical characteristics of study participants stratified by renal function.

| Characteristics | Overall | non-CKD | mild-CKD | MS-CKD | P-value |
| --- | --- | --- | --- | --- | --- |
| No. of subjects | 1408 | 119 | 1153 | 55 | - |
| Age, year | 64.4±7.8 | 59.1±8.9 | 64.7±7.5 | 71.3±5.6 | **<0.001** |
| Female, % | 774(58.33%) | 70(58.82%) | 671(58.2%) | 33(60.0%) | 0.959 |
| Duration of diabetes, year | 8.9±7.0 | 7.0±5.3 | 8.9±7.0 | 12.7±8.2 | **<0.001** |
| HbA1c, % | 7.0±1.5 | 7.0±1.5 | 6.9±1.4 | 7.0±1.5 | 0.857 |
| Body mass index, kg/m<sup>2</sup> | 24.6±3.3 | 24.6±3.6 | 24.5±3.2 | 25.2±3.1 | 0.269 |
| Systolic BP, mm Hg | 135.2±19.2 | 133.7±16.8 | 135.1±19.6 | 138.7±19.5 | 0.282 |
| Diastolic BP, mm Hg | 70.7±10.6 | 70.8±9.5 | 70.7±10.6 | 70.0±10.4 | 0.872 |
| Total cholesterol, mmol/L | 4.8±1.0 | 4.8±1.0 | 4.8±1.0 | 5.0±1.2 | 0.474 |
| Serum creatinine, µmol/L | 71.6±20.2 | 46.5±9.7 | 71.8±16.0 | 121.5±24.5 | **<0.001** |
| HDL-c, mmol/L | 1.3±0.4 | 1.3±0.4 | 1.3±0.4 | 1.2±0.4 | 0.134 |
| LDL-c, mmol/L | 3.1±0.9 | 3.0±0.9 | 3.1±0.9 | 3.2±1.1 | 0.464 |
| Triglycerides, mmol/L | 2.3±1.6 | 2.7±2.2 | 2.3±1.5 | 2.5±1.7 | **0.015** |
| Microalbuminuria, mg/mL | 5.3±17.4 | 1.9±5.6 | 5.3±17.7 | 16.1±29.2 | **<0.001** |
| BCVA, logMAR | 0.2±0.1 | 0.2±0.1 | 0.2±0.1 | 0.3±0.1 | **0.004** |
| Intraocular pressure, mmHg | 16.2±2.8 | 16.5±2.7 | 16.2±2.8 | 15.4±3.1 | 0.063 |
| Axial length, mm | 23.5±1.1 | 23.5±1.0 | 23.5±1.1 | 23.6±1.0 | 0.924 |
| DR status | 1058(79.73%) | 100(84.03%) | 918(79.62%) | 40(72.73%) | 0.219 |

CKD were categorized by using the eGFR cutoff of ≥90 for non-CKD, 90 to 60 for mild-CKD, 60-30 for moderate to severe CKD.
eGFR=estimated glomerular filtration rate; BP = Blood Pressure; HDL-C = High density lipoprotein-cholesterol; LDL-C = High density lipoprotein-cholesterol; BCVA = Best Corrected Visual Acuity; DR=diabetic retinopathy.
Bold indicates statistically significant

### Peripapillary CT and RNFL thickness in different groups

Table 2 shows the pCT and pRNFL thickness in different groups. The average pRNFL thickness also differed significantly among the three groups (114.0 μm ± 13.4 μm vs. 110.5 μm ± 12.8 μm vs. 107.4 μm ± 15.4 μm, P = 0.003). When analysing by quadrants, the pRNFL thickness was found to be superior and the nasal quadrants remained significant. Figure 2 shows the stratified analysis of the average pRNFL thickness by DR status.

**Table 2.** Peripapillary retinal nerve fiber layer (pRNFL) thickness and choroidal thickness (pCT) in patients with diabetes mellitus stratified by renal function.

| Parameters | no-CKD | mild-CKD | MS-CKD | P-value |
| --- | --- | --- | --- | --- |
| **pRNFL thickness, $\mu\text{m}$** |  |  |  |  |
| C1 | 152.9±25.3 | 146.4±26.4 | 137.8±28.5 | **0.002** |
| C2 | 118.8±26.8 | 108.2±25.6 | 105.7±24.3 | **<0.001** |
| C3 | 79.3±17.5 | 74.5±16.0 | 76.7±17.7 | **0.007** |
| C4 | 87.3±24.6 | 80.7±21.2 | 77.3±19.8 | **0.003** |
| C5 | 156.2±36.4 | 147.4±33.7 | 131.3±35.5 | **<0.001** |
| C6 | 164.7±35.2 | 163.9±37.6 | 147.1±39.7 | **0.005** |
| C7 | 121.6±33.4 | 122.0±33.8 | 118.8±36.4 | 0.790 |
| C8 | 78.3±23.9 | 79.4±22.2 | 81.5±24.9 | 0.680 |
| C9 | 68.2±18.8 | 67.7±17.1 | 70.1±17.0 | 0.576 |
| C10 | 95.0±26.0 | 95.6±24.2 | 100.0±27.7 | 0.405 |
| C11 | 117.0±25.7 | 113.8±25.7 | 115.0±25.7 | 0.421 |
| C12 | 128.9±33.1 | 125.7±30.4 | 125.5±31.8 | 0.570 |
| Superior | 132.9±22.0 | 128.7±20.5 | 126.1±22.7 | 0.061 |
| Inferior | 147.5±24.0 | 144.4±25.1 | 132.5±28.9 | **<0.001** |
| Nasal | 95.1±20.6 | 87.8±18.1 | 86.7±18.0 | **<0.001** |
| Temporal | 80.5±20.4 | 80.9±18.4 | 83.8±20.9 | 0.503 |
| Average | 114.0±13.4 | 110.5±12.8 | 107.4±15.4 | **0.003** |
| **pCT, $\mu\text{m}$** |  |  |  |  |
| C1 | 142.2±67.6 | 125.3±61.2 | 75.3±29.9 | **<0.001** |
| C2 | 140.4±72.4 | 124.1±66.9 | 72.2±36.1 | **<0.001** |
| C3 | 133.6±77.5 | 117.6±68.9 | 69.1±35.0 | **<0.001** |
| C4 | 118.5±69.8 | 105.8±62.1 | 64.5±28.4 | **<0.001** |
| C5 | 102.0±55.4 | 90.9±50.8 | 58.3±26.0 | **<0.001** |
| C6 | 91.8±46.7 | 83.2±44.3 | 54.1±23.0 | **<0.001** |
| C7 | 106.1±55.0 | 95.1±47.3 | 63.4±24.9 | **<0.001** |
| C8 | 122.7±62.8 | 109.3±53.0 | 73.1±25.9 | **<0.001** |
| C9 | 132.2±63.7 | 117.7±54.6 | 79.4±26.7 | **<0.001** |
| C10 | 136.9±64.8 | 123.8±56.5 | 81.5±34.0 | **<0.001** |
| C11 | 142.8±64.4 | 125.8±57.0 | 80.9±31.4 | **<0.001** |
| C12 | 142.4±61.5 | 125.1±58.2 | 79.7±30.4 | **<0.001** |
| Superior | 142.5±62.5 | 125.4±56.9 | 78.6±28.0 | **<0.001** |
| Inferior | 100.0±50.4 | 89.7±45.5 | 58.6±22.8 | **<0.001** |
| Nasal | 130.8±71.7 | 115.8±64.3 | 68.6±31.3 | **<0.001** |
| Temporal | 130.6±62.4 | 116.9±53.0 | 77.9±27.2 | **<0.001** |
| Average | 126.0±58.0 | 112.0±51.2 | 71.0±22.9 | **<0.001** |

eGFR=estimated glomerular filtration rate. CKD were categorized by using the eGFR cutoff of $\geq 90$ for non-CKD, 90 to 60 for mild-CKD, 60-30 for moderate to severe CKD (MS-CKD).
Bold indicates statistically significant.

**Figure 1.**
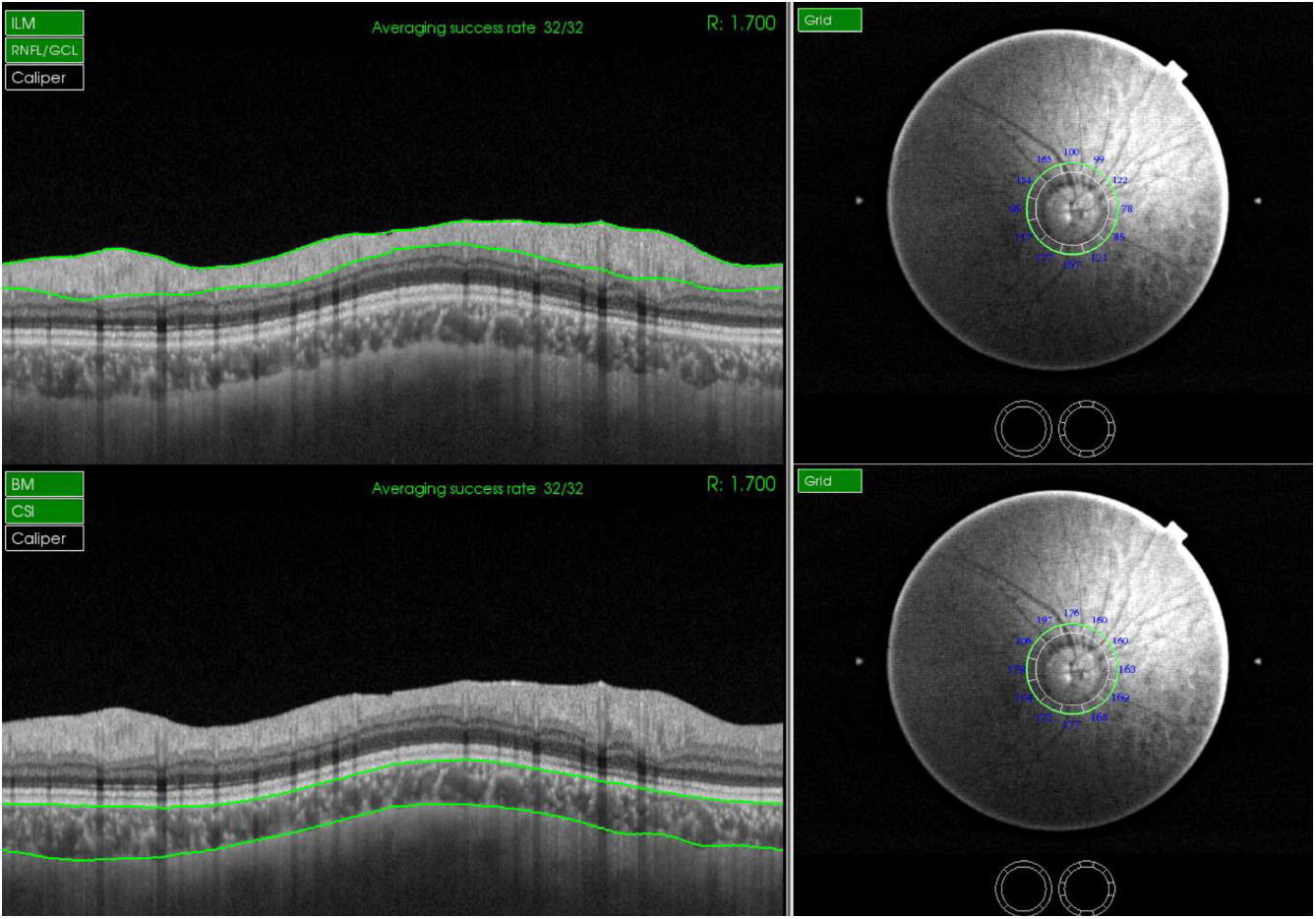
Measurement illustration of peripapillary retinal nerve fibre layer (pRNFL, A) thickness and choroidal thickness (pCT, B) displayed in 12 sectors of treatment-naïve patients with diabetes mellitus.

**Figure 2.**
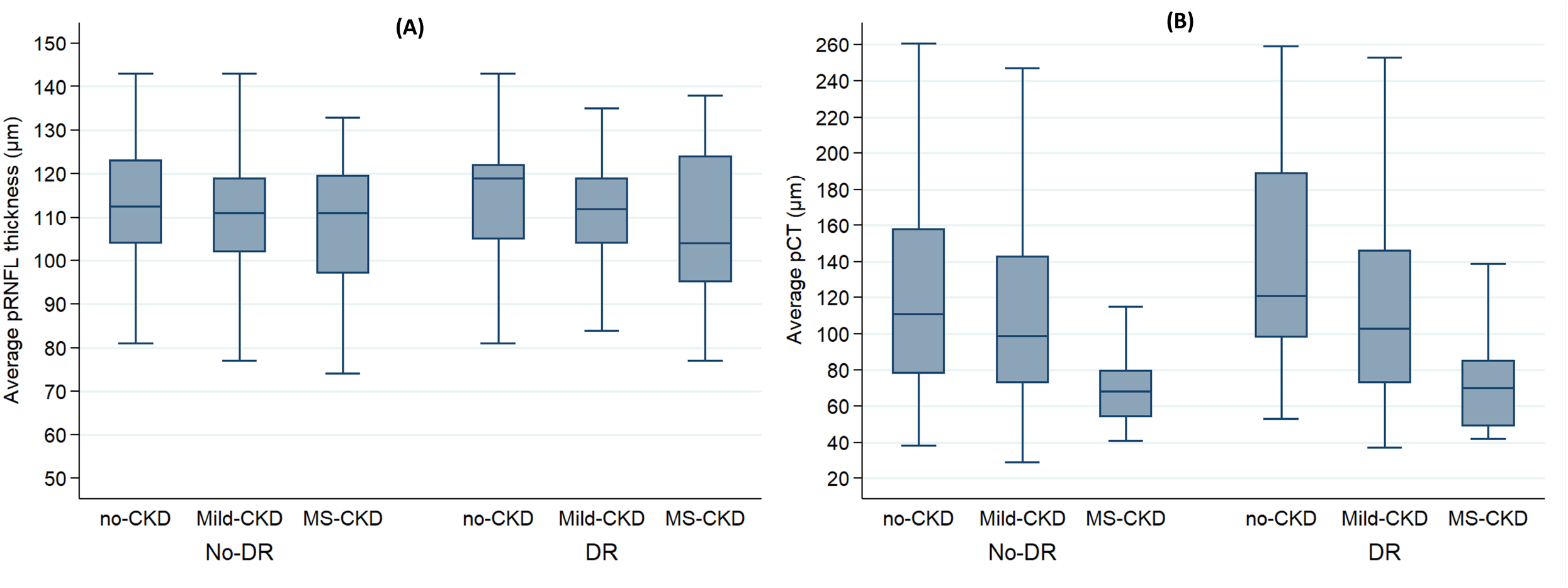
Box plots of average peripapillary retinal nerve fibre layer (pRNFL) thickness and choroidal thickness (pCT) in patients with and without diabetic retinopathy stratified by renal function. (A) pRNFL thickness; (B) pCT.

**Figure 3.**
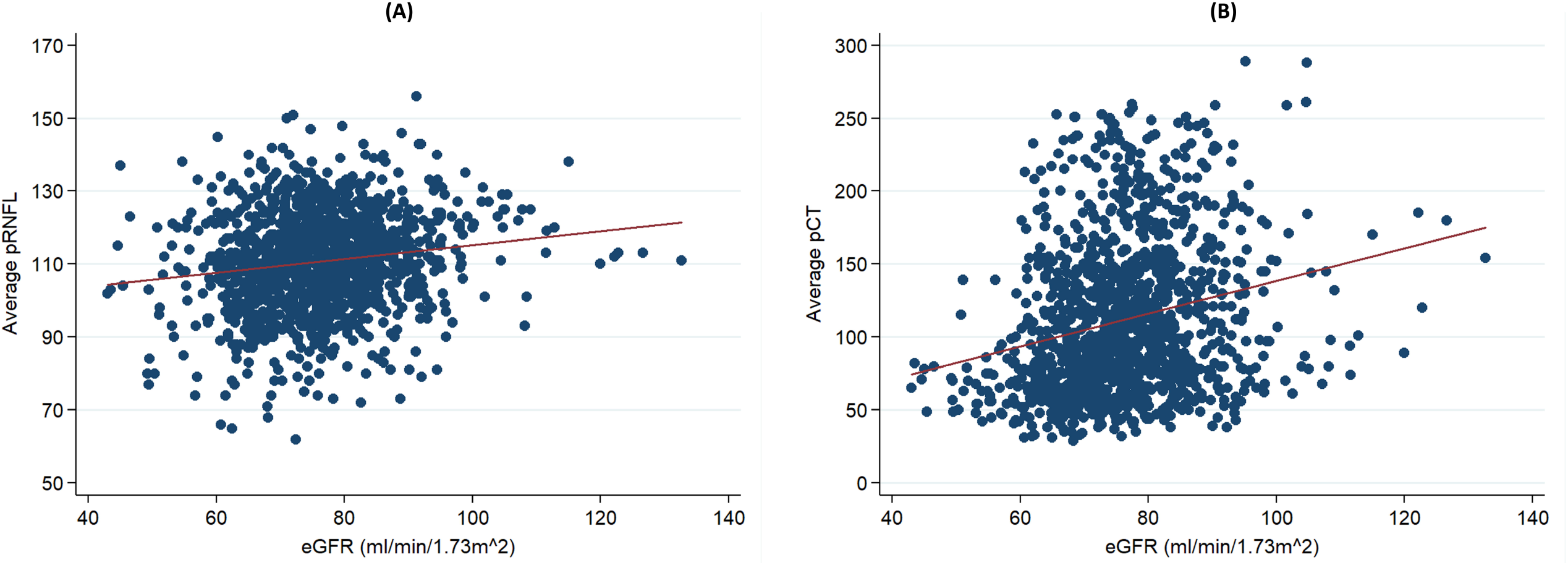
Scatter plots showing correlations of average pRNFL thickness and pCT with eGFR. (A) pRNFL versus eGFR; (B) pCT versus eGFR. eGFR=estimated glomerular filtration rate; pRNFL=peripapillary retinal nerve fibre layer; pCT= peripapillary choroidal thickness (pCT)

The mean pCT was significantly different among the three groups (126.0 ± 58.0 vs. 112.0 ± 51.2 vs. 71.0 ± 22.9, P < 0.001). Specifically, pCT in all regions showed significant intergroup differences (all P < 0.001). The mean pCT decreased with renal function in patients with and without DR. As the same renal function, the mean pCT in the DR group was thicker than the non-DR group.

Table 3 shows the result of regression analysis for which the renal function was taken as categorical variable. After adjusting for other factors, only the nasal pRNFL was found to be significantly negatively correlated with renal function (mild-CKD vs non-CKD P < 0.001, MS-CKD vs non-CKD P = 0.018). No statistically significant correlation was found between the average pRNFL thickness and pRNFL thickness in other regions.

**Table 3.**
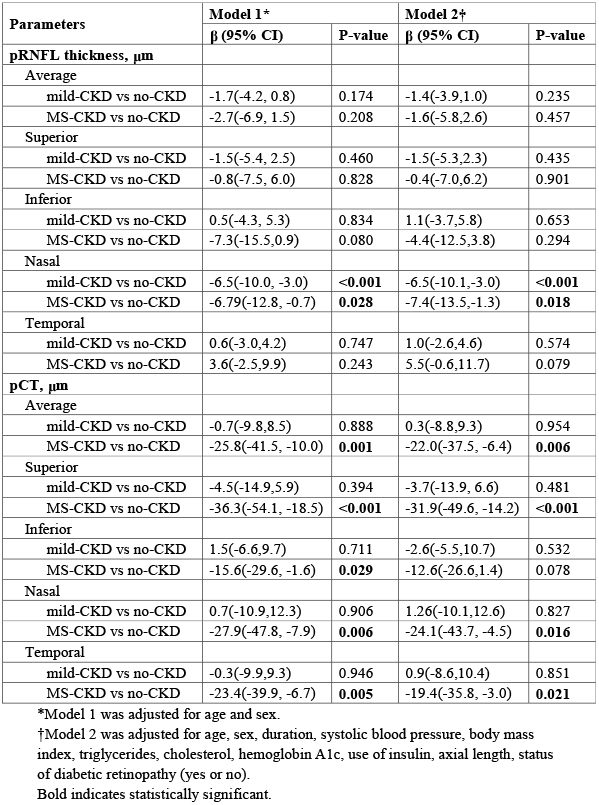
Comparisons of pRNFL thickness and pCT thickness between different groups after adjusting known confounding factors.

| Parameters | Model 1* |  | Model 2† |  |
| --- | --- | --- | --- | --- |
| | $\beta$ (95% CI) | P-value | $\beta$ (95% CI) | P-value |
| **pRNFL thickness, $\mu\text{m}$** |  |  |  |  |
| Average |  |  |  |  |
| mild-CKD vs no-CKD | -1.7(-4.2, 0.8) | 0.174 | -1.4(-3.9,1.0) | 0.235 |
| MS-CKD vs no-CKD | -2.7(-6.9, 1.5) | 0.208 | -1.6(-5.8,2.6) | 0.457 |
| Superior |  |  |  |  |
| mild-CKD vs no-CKD | -1.5(-5.4, 2.5) | 0.460 | -1.5(-5.3,2.3) | 0.435 |
| MS-CKD vs no-CKD | -0.8(-7.5, 6.0) | 0.828 | -0.4(-7.0,6.2) | 0.901 |
| Inferior |  |  |  |  |
| mild-CKD vs no-CKD | 0.5(-4.3, 5.3) | 0.834 | 1.1(-3.7,5.8) | 0.653 |
| MS-CKD vs no-CKD | -7.3(-15.5,0.9) | 0.080 | -4.4(-12.5,3.8) | 0.294 |
| Nasal |  |  |  |  |
| mild-CKD vs no-CKD | -6.5(-10.0, -3.0) | **<0.001** | -6.5(-10.1,-3.0) | **<0.001** |
| MS-CKD vs no-CKD | -6.79(-12.8, -0.7) | **0.028** | -7.4(-13.5,-1.3) | **0.018** |
| Temporal |  |  |  |  |
| mild-CKD vs no-CKD | 0.6(-3.0,4.2) | 0.747 | 1.0(-2.6,4.6) | 0.574 |
| MS-CKD vs no-CKD | 3.6(-2.5,9.9) | 0.243 | 5.5(-0.6,11.7) | 0.079 |
| **pCT, $\mu\text{m}$** |  |  |  |  |
| Average |  |  |  |  |
| mild-CKD vs no-CKD | -0.7(-9.8,8.5) | 0.888 | 0.3(-8.8,9.3) | 0.954 |
| MS-CKD vs no-CKD | -25.8(-41.5, -10.0) | **0.001** | -22.0(-37.5, -6.4) | **0.006** |
| Superior |  |  |  |  |
| mild-CKD vs no-CKD | -4.5(-14.9,5.9) | 0.394 | -3.7(-13.9, 6.6) | 0.481 |
| MS-CKD vs no-CKD | -36.3(-54.1, -18.5) | **<0.001** | -31.9(-49.6, -14.2) | **<0.001** |
| Inferior |  |  |  |  |
| mild-CKD vs no-CKD | 1.5(-6.6,9.7) | 0.711 | -2.6(-5.5,10.7) | 0.532 |
| MS-CKD vs no-CKD | -15.6(-29.6, -1.6) | **0.029** | -12.6(-26.6,1.4) | 0.078 |
| Nasal |  |  |  |  |
| mild-CKD vs no-CKD | 0.7(-10.9,12.3) | 0.906 | 1.26(-10.1,12.6) | 0.827 |
| MS-CKD vs no-CKD | -27.9(-47.8, -7.9) | **0.006** | -24.1(-43.7, -4.5) | **0.016** |
| Temporal |  |  |  |  |
| mild-CKD vs no-CKD | -0.3(-9.9,9.3) | 0.946 | 0.9(-8.6,10.4) | 0.851 |
| MS-CKD vs no-CKD | -23.4(-39.9, -6.7) | **0.005** | -19.4(-35.8, -3.0) | **0.021** |

\*Model 1 was adjusted for age and sex.
†Model 2 was adjusted for age, sex, duration, systolic blood pressure, body mass index, triglycerides, cholesterol, hemoglobin A1c, use of insulin, axial length, status of diabetic retinopathy (yes or no).
Bold indicates statistically significant.

**Table 4.** Univariable and multivariable regression analysis of the associations between pRNFL thickness/pCT and the eGFR.

| Per 1 eGFR increase | Model 1* |  | Model 2† |  |
| --- | --- | --- | --- | --- |
| | $\beta$ (95% CI) | P-value | $\beta$ (95% CI) | P-value |
| **pRNFL thickness, $\mu\text{m}$** |  |  |  |  |
| Average | 0.1(0.0,0.2) | **0.005** | 0.1(0.0,0.2) | **0.009** |
| Superior | 0.1(0.0,0.2) | 0.202 | 0.1(-0.1,0.2) | 0.282 |
| Inferior | 0.2(0.1,0.4) | **0.002** | 0.2(0.0,0.3) | **0.008** |
| Nasal | 0.2(0.1,0.3) | **0.002** | 0.2(0.1,0.3) | **<0.001** |
| Temporal | 0.0(-0.2,0.1) | 0.387 | -0.1(-0.2,0.0) | 0.243 |
| **pCT, $\mu\text{m}$** |  |  |  |  |
| Average | 0.4(0.2,0.7) | **0.002** | 0.3(0.0,0.6) | **0.021** |
| Superior | 0.6(0.3,0.9) | **<0.001** | 0.5(0.2,0.8) | **0.002** |
| Inferior | 0.3(0.0,0.5) | **0.030** | 0.2(-0.1,0.4) | 0.163 |
| Nasal | 0.4(0.1,0.8) | **0.014** | 0.3(0.0,0.6) | 0.077 |
| Temporal | 0.4(0.1,0.7) | **0.004** | 0.3(0.0,0.6) | **0.035** |

\*Model 1 was adjusted for age and sex.
†Model 2 was adjusted for age, sex, duration, systolic blood pressure, body mass index, triglycerides, cholesterol, hemoglobin A1c, use of insulin, axial length, status of diabetic retinopathy status (yes or no).
Bold indicates statistically significant.

In the analysis of pCT, the average pCT only showed a significant negative correlation between MS-CKD and non-CKD groups (p = 0.001), with similar results in the upper (p < 0.001), lower (p = 0.029), nasal (p = 0.006) and temporal (p = 0.005) quadrants in Model 1. The mean pCT was also negatively correlated only between the MS-CKD and non-CKD groups (p = 0.006), and similar results were still shown in the superior (p < 0.001), nasal (p = 0.016) and temporal (p = 0.021) quadrants.

### Association of pCT and RNFL thickness with eGFR

Figure 4 shows the positive correlation between average pRNFL thickness and the eGFR. After adjusting for age and sex, the eGFR was positively associated with the average pRNFL thickness (β = 0.1, 95%CI = 0.0-0.2, p = 0.005) and pRNFL thickness in the inferior (β = 0.2, 95%CI = 0.1-0.4, p = 0.002) and nasal (β = 0.2, 95%CI = 0.1-0.3, p = 0.002) quadrants. After further adjusting for other confounding factors, the eGFR was still significantly correlated with average pRNFL thickness (β = 0.1, 95%CI = 0.0-0.2, p = 0.009) and the inferior (β = 0.2, 95%CI = 0.0-0.3, p = 0.008) and nasal (β = 0.2, 95%CI = 0.1-0.3, p < 0.001) quadrants.

Figure 5 shows the positive correlation between average pCT and eGFR. After adjusting for age and sex, the eGFR was positively correlated with average pCT (β = 0.4, 95%CI = 0.2-0.7, p = 0.002) and pCT in all quadrants. Persistent results were obtained after further adjusting for other confounding factors, with average pCT (β = 0.3, 95%CI = 0.0∼0.6, p = 0.021) and pCT in the superior (β = 0.5, 95%CI = 0.2-0.8, p = 0.002) and temporal (β = 0.3, 95%CI = 0.0-0.6, p = 0.004) quadrants.

## Discussion

To the best of our knowledge, this is the first study to investigate the association between renal function and pRNFL thickness and pCT based on SS-OCT in Chinese patients with DM. This study demonstrated that pCT was significantly reduced in diabetic patients with CKD, and the higher average pCT was independently correlated with higher eGFR values. Similarly, the pRNFL thickness was found to be significantly thinner in diabetic patients with CKD, but only RNFL thickness in the nasal quadrant was independently correlated with the eGFR. These findings suggest that the impaired neural and choroidal microcirculation were biomarkers associated with renal impairment in diabetic patients.

The RNFL is the innermost part of the retinal structure, whose thinning reflects the condition of diabetic optic neuropathy. RNFL reduction in patients with impaired renal function was consistent with previous studies.^9^ A cross-sectional study by Choi et al.^9^ analysed the frequency of pRNFL defects in 96 non-glaucomatous patients with type 2 diabetes and reported that the urinary albumin excretion rate was significantly higher in patients with RNFL defects than in those without defects, and the urinary albumin-to-creatinine ratio was the only factor associated with visual field defect location after adjusting for other factors. The present study found that pRNFL thinning primarily occurred in the nasal quadrant. The inferior and temporal quadrants were reportedly most affected in glaucoma patients. The mechanisms of RNFL thinning caused by DN may differ from those associated with glaucoma. Given that the RNFL does not accept blood from the choroid, the thinning of the pRNFL may not be secondary to the thinning of the choroid, and the pathogenesis of pRNFL and choroid thinning may be different.^15, 16^

The choroid is a highly vascularized structure that plays an important role in the regulation of ocular metabolism. Previous studies on CT and renal function have primarily focused on the macular area.^10-13, 17, 18^ Previous studies on macular choroid in non-diabetic population found a correlation between eGFR and CT to some extent.^17, 18^ More attention should be paid to the peripapillary choroid because one of the major functions of peripapillary choroid is to supply blood to the anterior layer of optic nerve head, and peripapillary atrophic changes are related to optic nerve-related diseases.^19, 20^ This study expanded previous research involving the peripapillary choroid, indicating that renal function was not only strongly associated with macular CT, but also for pCT. The impaired renal function may lead to ischemia of the optic nerve by affecting pCT, which may contribute to the pathogenesis mechanism of diabetic optic neuropathy.

The pCT in patients with MS-CKD were significantly thinner than those in non-CKD, while the pCT in patients with mild-CKD were not statistically significantly higher than those in non-CKD patients. This suggests that pCT may not change significantly when renal function is slightly reduced, but is altered when renal function is severely reduced. We speculated that the decline of renal function may occur before there is a change in the peripapillary structure, which needs to be further confirmed by longitudinal studies.

The underlying mechanism of renal function loss and pCT thinning remains unclear. Choroid, as a vascular structure, thinning reflects microvascular injury, which may indicate that morphological changes in the choroid may reflect systemic vascular injury.^21, 22^ It was reported that renal impairment was associated with oxidative stress, subclinical inflammation, and endothelial dysfunction.^23^ Hence, these factors may explain why choroid thinning is commonly found in patients with early renal impairment. The microvascular endothelial dysfunction was associated with proteinuria, and this association was more obvious in patients with type 2 diabetes.^8^ It also reported that CT was negatively related to the biomarkers of endothelial dysfunction and vascular inflammation, such as endothelin-1, ADMA and IL-6.^17^ The choroid injury may also be secondary to retinopathy because an alteration of the outer retinal layer and the RPE are nourished by the choroid vessels. In addition, dysfunction of the autonomic nervous system in patients with renal impairment plays a role in choroid thinning.^5^ More studies are needed to ascertain the underling mechanism.

Many factors affect mCT and pCT, including age, sex, HbA1c, duration of DM, AL, BP, serum lipid and history of ocular treatment (such as retina laser photocoagulation, anti-VEGF treatment, intravitreous injection and hemodialysis).^21, 22^ This study only included ocular treatment-naïve patients, and adjustments were made for the potential confounding factors in statistical analyses. In addition, the innovative SS-OCT was adopted in this study. Because of limited image resolution and repeatability, a quantitative evaluation of the choroid has been a challenging task for traditional imaging methods. Histological evaluation is also difficult due to a lack of biological samples and alterations caused by tissue fixation. The introduction of enhanced depth imaging SD-OCT (EDI SD-OCT) makes it possible to obtain cross-sectional images of the choroid. However, EDI SD-OCT also has limitations, such as the presence of an unclear boundary of the choroid, errors due to manual measuring methods, and poor representation associated with single-point measurements.^24^ The SS-OCT has a higher scanning speed, higher resolution, clearer choroid-scleral interface and more accurate automatic measurements compared to EDI SD-OCT.

This study does have certain limitations. First, it is not possible to determine the causal relationship due the nature of this cross-sectional study. Secondly, the findings of the present study were only applicable to type 2 diabetes patients from the Chinese population. Future multi-centre studies with various ethnicities are needed to verify the conclusions. Thirdly, the sample size of the MS-CKD group was relatively small, which may lower the ability of the test to find statistical differences in this group. However, we still found that pCT/pRNFL thickness decreased with the deterioration of renal function. Finally, blood and urine samples were collected at a single point in time, so the calculated eGFR may not accurately represent the renal function level of the participants due to accidental factors, which may lead to nondifferential misclassification bias in CKD grouping.

In summary, SS-OCT, a novel imaging technology, was employed to find that pCT and pRNFL thickness both became thinner as renal function declined in the Chinese diabetic population, and pCT/pRNFL thickness was positively correlated with eGFR. These findings confirm the close relationship between alterations in ocular microcirculation and renal impairment. Further studies are needed to elucidate the pathogenic mechanism behind the findings and to clarify the role of CT in the diagnosis and prognosis of DR and DN in patients with type 2 diabetes.

## Data Availability

All data were in manuscript.

## Acknowledgement

This study was supported by the National Natural Science Foundation of China (81570843; 81530028; 81721003), the Guangdong Province Science & Technology Plan (2014B020228002).

## Author Contributions

WW and WH had full access to all the data in the study and take responsibility for the integrity of the data and the accuracy of the data analysis. *Study concept and design:* WW, WL, MH, WH.

*Acquisition, analysis, or interpretation of data:* LS, DZ, YT, YL, WL.

*Drafting of the manuscript:* LS, DZ, WW.

*Critical revision of the manuscript for important intellectual content:* All authors.

*Statistical analysis:* WW.

*Obtained funding:* WH.

*Administrative, technical, or material support:* MH, WW.

*Study supervision:* MH.

## Data sharing statement

The data used and/or analyzed during the current study are available from the corresponding author on reasonable request.

## References

1. NCD Risk Factor Collaboration (NCD-RisC). Worldwide trends in diabetes since 1980: A pooled analysis of 751 population-based studies with 4.4 million participants. Lancet 2016;387:1513–30.

2. Valencia WM, Florez H. How to prevent the microvascular complications of type 2 diabetes beyond glucose control. BMJ 2017;356:i6505.

3. Elias MF, Torres RV, Davey A. The eye is the window to the kidney and brain. Ebiomedicine 2016;5:24–5.

4. Vistisen D, Andersen GS, Hulman A, Persson F, Rossing P, Jorgensen ME. Progressive decline in estimated glomerular filtration rate in patients with diabetes after moderate loss in kidney Function-Even without albuminuria. Diabetes Care 2019;42:1886–94.

5. Wong CW, Wong TY, Cheng CY, Sabanayagam C. Kidney and eye diseases: Common risk factors, etiological mechanisms, and pathways. Kidney Int 2014;85:1290–302.

6. Wong TY, Coresh J, Klein R, Muntner P, Couper DJ, Sharrett AR, Klein BE, Heiss G, Hubbard LD, Duncan BB. Retinal microvascular abnormalities and renal dysfunction: The atherosclerosis risk in communities study. J Am Soc Nephrol 2004;15:2469–76.

7. Sabanayagam C, Shankar A, Klein BE, Lee KE, Muntner P, Nieto FJ, Tsai MY, Cruickshanks KJ, Schubert CR, Brazy PC, et al. Bidirectional association of retinal vessel diameters and estimated GFR decline: The Beaver Dam CKD Study. Am J Kidney Dis 2011;57:682–91.

8. Martens R, Houben A, Kooman JP, Berendschot T, Dagnelie PC, van der Kallen C, Kroon AA, Leunissen K, van der Sande FM, Schaper NC, et al. Microvascular endothelial dysfunction is associated with albuminuria: The Maastricht Study. J Hypertens 2018;36:1178–87.

9. Choi JA, Ko SH, Park YR, Jee DH, Ko SH, Park CK. Retinal nerve fiber layer loss is associated with urinary albumin excretion in patients with type 2 diabetes.Ophthalmology 2015;122:976–81.

10. Kocasarac C, Yigit Y, Sengul E, Sakalar YB. Choroidal thickness alterations in diabetic nephropathy patients with early or no diabetic retinopathy. Int Ophthalmol 2018;38:721–6.

11. Garrido-Hermosilla AM, Mendez-Muros M, Gutierrez-Sanchez E, Morales-Portillo C, Diaz-Granda MJ, Esteban-Gonzalez E, Relimpio-Lopez I, Martinez-Brocca MA, Rodriguez-de-la-Rua-Franch E. Renal function and choroidal thickness using swept-source optical coherence tomography in diabetic patients. Int J Ophthalmol 2019;12:985–9.

12. Farias LB, Lavinsky D, Schneider WM, Guimaraes L, Lavinsky J, Canani LH. Choroidal thickness in patients with diabetes and microalbuminuria. Ophthalmology 2014;121:2071–3.

13. Malerbi FK, Regatieri CV, de Sa JR, Morales PH, Farah ME, Dib SA. Microalbuminuria is associated with increased choroidal thickness in type 1 diabetes mellitus patients without diabetic retinopathy. Acta Ophthalmol 2018;96:e95–7.

14. Li DY, Yin WJ, Yi YH, Zhang BK, Zhao J, Zhu CN, Ma RR, Zhou LY, Xie YL, Wang JL, et al. Development and validation of a more accurate estimating equation for glomerular filtration rate in a Chinese population. Kidney Int 2019;95:636–46.

15. Gupta P, Cheung CY, Baskaran M, Tian J, Marziliano P, Lamoureux EL, Cheung CM, Aung T, Wong TY, Cheng CY. Relationship between peripapillary choroid and retinal nerve fiber layer thickness in a Population-Based sample of nonglaucomatous eyes. Am J Ophthalmol 2016;161:4–11.

16. Lin S, Cheng H, Zhang S, Ye C, Pan X, Tao A, Xu X, Qu J, Liang Y. Parapapillary choroidal microvasculature dropout is associated with the decrease in retinal nerve fiber layer thickness: A prospective study. Invest Ophthalmol Vis Sci 2019;60:838–42.

17. Balmforth C, van Bragt JJ, Ruijs T, Cameron JR, Kimmitt R, Moorhouse R, Czopek A, Hu MK, Gallacher PJ, Dear JW, et al. Chorioretinal thinning in chronic kidney disease links to inflammation and endothelial dysfunction. JCI Insight 2016;1:e89173.

18. Mule G, Vadala M, La Blasca T, Gaetani R, Virone G, Guarneri M, Castellucci M, Guarrasi G, Terrasi M, Cottone S. Association between early-stage chronic kidney disease and reduced choroidal thickness in essential hypertensive patients. Hypertens Res 2019.

19. Ho J, Branchini L, Regatieri C, Krishnan C, Fujimoto JG, Duker JS. Analysis of normal peripapillary choroidal thickness via spectral domain optical coherence tomography. Ophthalmology 2011;118:2001–7.

20. Lee SH, Lee EJ, Kim TW. Topographic correlation between juxtapapillary choroidal thickness and microstructure of parapapillary atrophy. Ophthalmology 2016;123:1965–73.

21. Ferrara D, Waheed NK, Duker JS. Investigating the choriocapillaris and choroidal vasculature with new optical coherence tomography technologies. Prog Retin Eye Res 2016;52:130–55.

22. Tan KA, Gupta P, Agarwal A, Chhablani J, Cheng CY, Keane PA, Agrawal R. State of science: Choroidal thickness and systemic health. Surv Ophthalmol 2016;61:566–81.

23. Anders HJ, Huber TB, Isermann B, Schiffer M. CKD in diabetes: Diabetic kidney disease versus nondiabetic kidney disease. Nat Rev Nephrol 2018;14:361–77.

24. Singh SR, Vupparaboina KK, Goud A, Dansingani KK, Chhablani J. Choroidal imaging biomarkers. Surv Ophthalmol 2019;64:312–33.

